# Cross-infection of *Ascaris* spp. in humans and pigs from a Guarani indigenous village in southern Brazil

**DOI:** 10.1101/2024.06.07.24308590

**Authors:** Veridiana Lenartovicz Boeira, Renata Coltro Bezagio, Marina Silva de Carvalho, Rinaldo Ferreira Gandra, Ana Paula de Abreu, Cristiano Lara Massara, Cristiane Maria Colli, Max Jean de Ornelas Toledo

**Affiliations:** Postgraduate Program in Biological Sciences, State University of Maringá, Maringá, Paraná, Brazil; Pharmacy Course, State University of Western Paraná, Cascavel, Paraná, Brazil; Postgraduate Program in Health Sciences, State University of Maringá, Maringá, Paraná, Brazil; Helminthology Laboratory, René Rachou Institute, FIOCRUZ, Belo Horizonte, Minas Gerais, Brazil; Federal University of Grande Dourados, Dourados, Mato Grosso do Sul, Brazil

**Author notes:** Corresponding author: (MJOT).

## Abstract

**Background:** *Ascaris lumbricoides* and *Ascaris suum* are nematode parasites that infect millions of people and pigs worldwide, respectively. Reports of cross-infection between the two host species has stimulated molecular epidemiological studies of the *Ascaris* genus. In this study, we evaluated the dynamics of *Ascaris* transmission between Guarani indigenous schoolchildren, pigs, and the environment of a village in the state of Paraná, southern Brazil.

**Methodology/Principal findings:** Parasitological and molecular analyses of fecal samples from humans and pigs, and soil samples from the village were carried out. Eggs of *Ascaris* spp. were observed in 8.4% (7/83) of human samples, 44.4% (8/18) of pig samples, and 8.9% (6/68) of soil samples. PCR amplification of the *ITS-1* locus of the rDNA gene in samples that were positive in the parasitological examination revealed cross-infection by the two species, *A. lumbricoides* and *A. suum*, in human and swine hosts. The soil, which was contaminated by both human and swine feces, also contained eggs of the two *Ascaris* species, thus constituting a source of *Ascaris* infection for both hosts. DNA from both nematode species, individually and mixed, was detected in samples from both hosts and the soil.

**Conclusions/Significance:** The results of this study indicate that more effective control measures, aimed at the correct disposal of both human and animal feces, should be implemented.

**Author Summary:** Despite the control measures implemented in Brazilian Indigenous Lands, the prevalence of intestinal parasites continues to vary from moderate to high in its inhabitants. On the other hand, the number of indigenous people residing in Brazil has been increasing in last decades, including the South region of the country, where a much smaller proportion of indigenous people reside compared to the North region. *Ascaris lumbricoides* and *Ascaris suum* are the most prevalent helminth parasites in humans and pigs, respectively. The possibility of cross-infection by *Ascaris* spp. between humans and pigs has been analyzed in order to propose more effective control measures. In this research, we use parasitological and molecular methods to verify the presence of these parasites in fecal samples from schoolchildren and pigs, in addition to soil samples, from a Guarani indigenous village in the state of Paraná, southern Brazil. Genetic material from both individual and associated nematode species was detected in host and soil samples, indicating cross-transmission in these populations. Control measures aimed at the correct disposal of human and animal feces must be implemented in order to minimize damage to health and prevent new infections.

## Introduction

*Ascaris lumbricoides* is the most prevalent geohelminth in regions endemic for soil-transmitted helminths. This nematode parasite is a cosmopolitan with wide geographic distribution; around 500 million individuals are infected worldwide and infection rates are above 20% in certain regions of Asia, sub-Saharan Africa, and Latin America [1–3]. In these regions, the high prevalence is closely related to the lack of basic sanitation and poor hygiene practices, conditions directly associated with poverty, in addition to climatic factors that favor the biological cycle of the parasite [4]. Ascariasis is therefore considered a neglected tropical disease and, although the majority of cases do not lead to death, this disease causes approximately 604 thousand years of life lived with disability, representing a serious economic and health problem in the affected countries [5,2].

The estimated percentage of pig herds infected by intestinal parasites worldwide is high (∼50%), particularly among pigs raised in extensive and organic animal production systems when compared to those in industrialized systems [6–7]. One of the most reported species of parasite in pig herds is the helminth *Ascaris suum*, which was first described by Goeze in 1782 [6–9].

*Ascaris lumbricoides* and *Ascaris suum* are considered cryptic because they have great biological and morphological similarity; the species can only be differentiated by differences in the labial denticles of adult helminths using scanning electron microscopy. Species-specific identification through visualization of eggs by optical microscopy is impossible, since they are morphologically identical [10–12].

The real scenario of human and swine ascariasis, in endemic and non-endemic regions, is still unknown and is difficult to address by conventional coproparasitological methods, such as microscopy. Comparative analyses of ribosomal DNA (rDNA) comprising the internal transcribed spacer 1 (*ITS-1*) locus revealed deletions in the *A. lumbricoides* sequence and polymorphisms in the alignment between the two species, totaling six divergent nucleotide positions. The sum of these factors results in a genetic divergence of 1.3% between the *ITS-1* sequences of *A. lumbricoides* and *A. suum*, with a variation of up to 0.2% within the taxon, enabling the species to be differentiated by molecular techniques [13–15].

According to 2022 census data, the number of indigenous people residing in Brazil was 1,693,535, which represented 0.83% of the country’s total population; this was an 88.82% increase from the 2010 census number [16]. More than half of Brazilian indigenous people live in the Legal Amazon while 44.48% (753,284 people) are concentrated in the North region of the country [16]. Only 5.20% of this population (88,097 people) reside in the South region of Brazil.

The prevalence of enteroparasitosis in the Brazilian indigenous population continues to be moderate to high, despite the implementation of control activities, such as housing improvements, water pipe installation and water treatment, and mass antiparasitic treatment of the population [17–19]. Therefore, reducing the morbidity and mortality caused by enteroparasite infection in this population still remains a challenge for health authorities.

In principle, indigenous people have more frequent contact with domestic animals and, to a certain extent, with wild animals, than the rest of society. The link between pathogens, the environment, animal species, and humans is inseparable, and thus, developing strategies to avoid or minimize current health problems requires collaboration from different fields of research. This approach is called “One Health” [20] and, following the COVID-19 pandemic, recognition of its relevance has grown [21].

In this study, using a One Health approach, we sought to analyze the dynamics of transmission of *Ascaris* spp. in a Guarani indigenous village in Paraná, southern Brazil, through the identification and molecular characterization of isolates obtained from samples of human and swine feces and soil.

## Materials and methods

### Ethical aspects

This study was approved by the National Research Ethics Commission through Opinion 1.756.060/2016, with participation in the study conditional on the signing of the free and informed consent form by the parents/guardians of the schoolchildren that provided fecal samples. The research was authorized by local indigenous leaders and the Special Indigenous Health District - South Coast.

### Study area

In the state of Paraná, located in the South region of Brazil, 23 indigenous lands/villages exist, corresponding to an area of 85,264.30 hectares [22]. The Tekoha Ocoy Indigenous Land, called Santa Rosa do Ocoy village, is located in the municipality of São Miguel do Iguaçu, west of Paraná (25° 20′ 50″ S; 54° 14′ 6″ W). The village is located 14 km away from the urban center of São Miguel do Iguaçu and covers a territory of around 250 hectares (Fig 1). The village receives assistance from the National Indigenous People Foundation, the National Health Foundation, the Itaipu Binacional Hydroelectric Plant, and the São Miguel do Iguaçu City Hall. The people of the village use Portuguese and Guarani as a form of communication.

**Fig 1.**
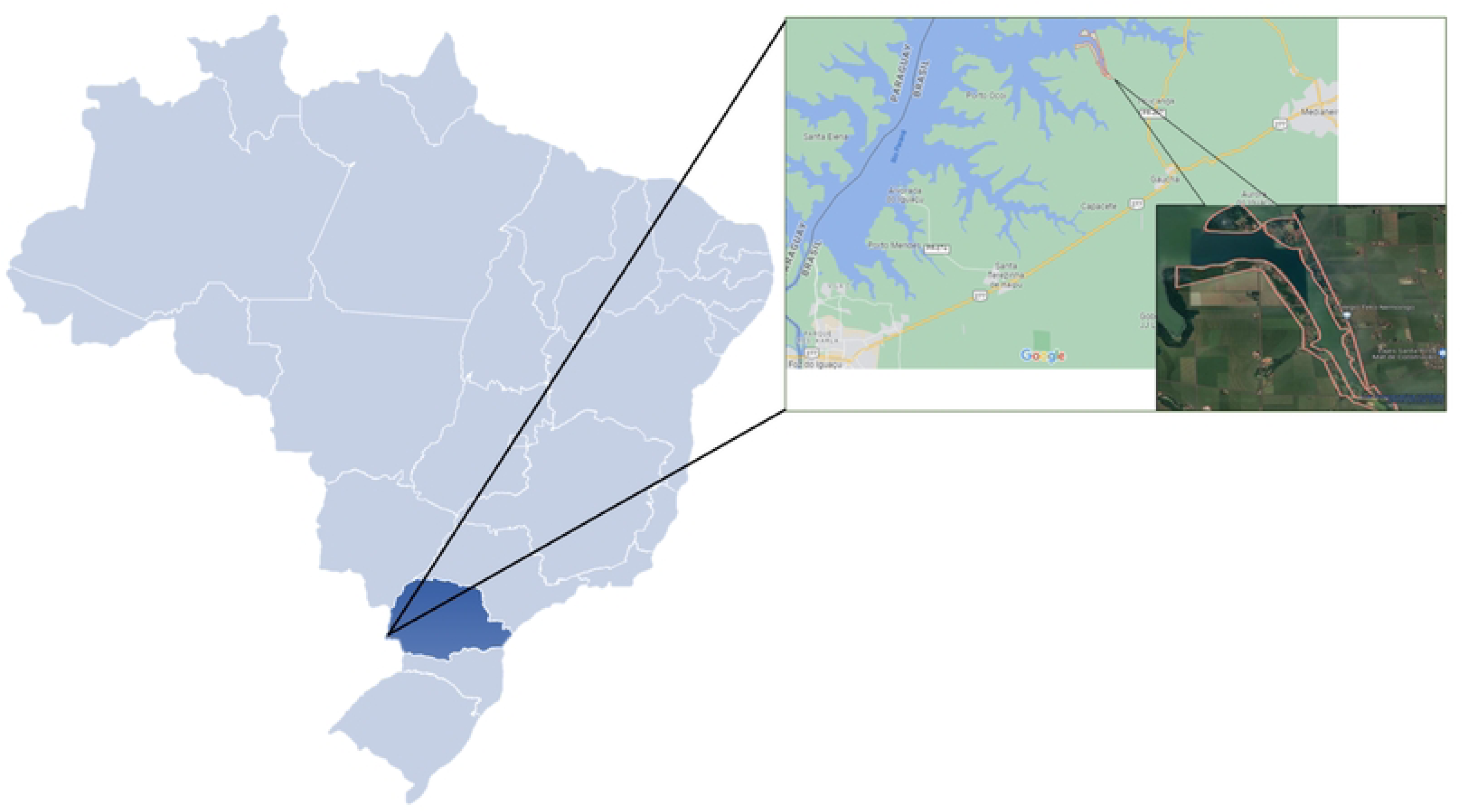
Map showing the geographic location and physical delimitation of Santa Rosa do Ocoy village of the Guarani indigenous people, located in municipality of São Miguel do Iguaçu, Paraná, Brazil. Shaded area of the map of Brazil represents the state of Paraná. within this area indicates the indigenous land. Adapted from the Central Intelligence Agency (CIA – USA). Source: the author

The indigenous community is located on the edge of Itaipu lake, with the residences surrounding the lake, divided into groups according to the kinship between the families. The houses are made of stone and wood and have straw-thatched roofs. In addition, the village has community spaces, including a prayer house, football fields, a warehouse for meetings and gatherings, a school, and a basic health unit [23].

### Population

According to the 2022 census, approximately 30,460 indigenous people were living in Paraná [16], belonging to three ethnicities: Guarani, Kaingáng, and Xetá. The village of Santa Rosa do Ocoy is inhabited by approximately 500 people belonging to the Guarani ethnic group, who are distributed among 106 households. Around 50% of the population is under 15 years of age, which is a common characteristic of the indigenous peoples of Brazil [16].

The study population consisted of schoolchildren, aged between 5 and 19 years, following the Toledo distribution [24]. All students enrolled at the local indigenous school, Colégio Indígena Teko Nemoingo, from nursery to high school age, were invited to participate in the study (around 270 students).

### Sample collection

Collection bottles (polypropylene bottles with a screw cap) for fecal samples, duly labeled with the participants’ names, were delivered, along with verbal and written instructions on how to perform the sample collection, to all children at the school, with the help of indigenous community health agents. The sample collection was carried out by parents, guardians, or by the children themselves if they were able to understand and correctly carry out the collection on their own, between November 2019 and August 2022. The bottles containing the samples were collected by the village health team and placed in thermal boxes with reusable artificial ice, which were sent to the Clinical Parasitology Laboratory of the University of Western Paraná (UNIOESTE), located in the Teaching, Research and Extension Laboratory of the University Hospital of Western Paraná, within 24 h of collection.

Samples of pig feces were collected randomly from fresh evacuations observed by the research team in the closed pigsties. The samples were placed in duly identified collection bottles, which were then placed in thermal boxes with artificial ice and sent to the same laboratory for analysis.

For the soil samples, collection points were chosen close to the school, residences with animals raised in the home, and residences in different locations within the village area, respecting a perimeter of 10 m around each collection location. Approximately 50 g of soil were collected from each point at a depth of 5 cm. Different climatic and humidity conditions that could influence the results were taken into account, and thus, collection was repeated at the same points in each of the four seasons of the years 2019 and 2021. The samples were placed in collection bottles, labelled with the date and location of collection, and then sent for analysis in the same way as the fecal samples. No preservatives were used for any type of sample collected.

### Soil sample treatment

The soil of the village was classified as clayey, making it difficult to extract compounds as it is more compacted than other types of soil [25]. To reduce dirt and interference in the analyses, the concentration and purification protocol of the Ministry of Health was performed [26], where 10 g of sample was mixed with 1 M glycine to a volume of 40 mL, homogenized for 30 min at 20 rpm, then completed to 50 mL with 1 M glycine and left to rest for 5 min. The supernatant was then transferred to another tube, centrifuged at 2,100 × g for 10 min, and the sediment was subjected to parasitological analysis.

### Parasitological analyses

Analysis of the samples was carried out at the UNIOESTE Clinical Parasitology Laboratory. When immediate analysis was not possible, the samples were kept refrigerated for a maximum of 2 days.

Fecal samples from humans (n=86), together with those from pigs (n=18) and soil (n=68) were processed by spontaneous sedimentation in water [27]. The sediments (3 mL) were then subjected to the Ritchie method [28] adapted by Bezagio et al. [29].

The samples were placed on slides and stained with Lugol, which were then examined by optical microscopy on an Olympus CX31^®^ microscope (Olympus Corporation, Tokyo, Japan) with a 10× objective; structures were confirmed at 400× magnification. The presence of parasite eggs, larvae, or cysts was considered a positive result, regardless of the quantity.

### Molecular analyses

Molecular analyses for the detection of *Ascaris* genetic material and identification of the species was carried out for all fecal and soil samples, regardless of the microscopy results. Positive and negative controls were also used.

### DNA extraction

First, the efficiency of two commercial DNA extraction kits was tested using samples from the laboratory that were known to be positive for *Ascaris* spp. The PureLink PCR Purification^®^ Kit (Invitrogen, Carlsbad, CA, USA) was used for laboratory samples that had or had not undergone prior sonication treatment for disruption of the *Ascaris* spp. egg membrane: 50 Hz for 30 s at 4°C, repeated 4 times, with an interval of 1 min between cycles. The QIAmp^®^ DNA Stool Mini Kit (Qiagen, Hilden, Germany), which is widely used for DNA extraction from fecal samples, was used only for laboratory samples that had not undergone the sonication procedure. The intensity of the bands observed in an electrophoresis gel after PCR with the extracted samples informed which DNA extraction method was subsequently used for the collected samples.

### Amplification of the *ITS-1* fragment

The approximately 580 base pair (bp) fragment of the *ITS-1* locus was amplified by nested PCR with the primers F2662 5′- GGCAAAAGTCGTAACAAGGT-3′ and R3214 5′- CTGCAATTCGCACTATTTATCG-3′, according to Ishiwata et al. [30]. Each amplification reaction was performed in a final volume of 10 μL, containing 1× reaction buffer (200 mmol/L Tris-HCl pH 8.4, 500 mmol/L KCl), 2.5 mmol/L MgCl_2_, 1 U of Platinum^®^ Taq DNA Polymerase (Invitrogen, Lithuania), 200 μmol/L deoxyribonucleotide triphosphate, 2 pmol of each primer, sterile Milli-Q^®^ water, and 2 μL of DNA. Amplification conditions were used according to Sadaow et al. [14]: pre-incubation at 95°C for 5 min, followed by 35 cycles at 95°C for 30 s, 55°C for 30 s, and 72°C for 30 s, and then a final extension at 72°C for 10 min.

### Genotyping of *Ascaris* spp

Three primers were used in the same reaction; two of which were species-specific forward primers, F. *Al* specific for *A. lumbricoides* (5′-GCGGTTTCTTTT TTTTTTCGCG-3′) and F. *As* specific for *A. suum* (5′-GAGAAAGCTCCTCGT TGCGG-3′), and a reverse primer (5′-CCACGAACCGAGTGATCCAC-3′), which anneals to a region common to both species (R. *both*). Amplification of genetic material from *A. lumbricoides* results in a fragment of approximately 384 bp, with the combination of primers F. *Al* + R. *both*, while for *A. suum*, the combination of primers F. *As* + R. *both* results in a fragment of approximately 176 bp. This technique allows either or both species to be detected in the same reaction (Fig 2). Conventional PCRs were performed, in a final volume of 10 μL, containing 1× reaction buffer (200 mmol/L Tris-HCl pH 8.4, 500 mmol/L KCl), 2.5 mmol/L MgCl_2_, 1 U of Platinum^®^ Taq DNA polymerase (Invitrogen, Lithuania), 200 μmol/L deoxyribonucleotide triphosphate, 2 pmol of each primer, sterile Milli-Q^®^ water, and 5 μL of DNA from the first reaction. Amplifications were performed using the following program: pre-incubation at 95°C for 5 min, 30 cycles at 95°C for 30 s, 60°C for 45 s, and 72°C for 45 s, and a final extension step at 72°C for 8 min [31].

**Fig 2.**
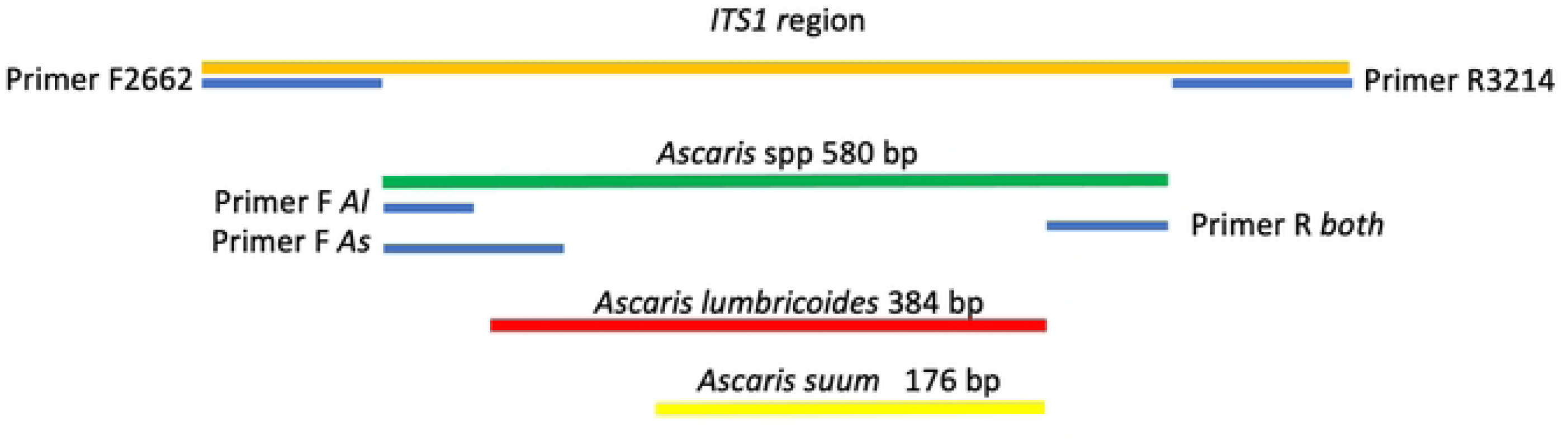
Molecular identification scheme of *Ascaris* spp. based on the *ITS-1* locus, using DNA extracted from the egg suspension. The first PCR used primers F2662 and R3214 for the amplification of the *ITS-1* locus (∼580 bp), and thus, the identification of *Ascaris* spp. The second PCR used the fragment resulting from the first PCR, together with two species-specific forward primers (F *Al* and F *As*) and a common reverse primer (R *both*), for the identification of *A. lumbricoides* (∼384 bp) and *A. suum* (∼176 bp). Source: the author

In all reactions, a positive control for *A. lumbricoides* was used (DNA provided by Helminthology Laboratory of the René Rachou Institute), a positive control for *A. suum* (DNA extracted from samples of adult worms from swine), and a negative control with Milli-Q^®^ water in place of DNA were used.

The products of these reactions were visualized on 5% polyacrylamide gels, stained with silver, and digitally photographed.

## Results

All students (approximately 270), aged between 5 and 19, enrolled in the indigenous school of the village were invited to participate in the study; however, only 86 bottles (approximately 30.5%) containing fecal samples were returned for analysis. Three samples were excluded because they had an insufficient amount of material to carry out the analyses. The total prevalence of intestinal parasites in the schoolchildren was 81.9% (68/83); 47.0% (39/83) had polyparasitism while the specific prevalence of *Ascaris* spp. was 8.4% (7/83) Table 1.

**Table 1.** Total and species-specific prevalence of intestinal protozoa and helminths in indigenous Guarani schoolchildren from the village of Santa Rosa do Ocoy, municipality of São Miguel do Iguaçu, state of Paraná, southern Brazil, between November 2019 and August 2022 (n=83).

|  | **No. positive samples** | **Prevalence (%)** |
| --- | --- | --- |
| **Total** | 68 | 81.9 |
| **Polyparasitism** | 39 | 47.0 |
| **Protozoa** | 62 | 74.7 |
| *Entamoeba coli* | 34 | 40.9 |
| *Endolimax nana* | 30 | 36.1 |
| *Giardia duodenalis* | 26 | 31.3 |
| *Entamoeba histolytica/E. dispar* | 21 | 25.3 |
| *Blastocystis hominis* | 8 | 9.6 |
| *Iodamoeba butschlii* | 6 | 7.2 |
| **Helminths** | 27 | 32.5 |
| *Hymenolepis nana* | 22 | 26.5 |
| *Ascaris* spp. | 7 | 8.4 |
| **Hookworms** | 4 | 4.8 |
| *Trichuris trichiura* | 1 | 1.2 |

Fecal samples from pigs were collected in March 2020, which coincided with the beginning of the COVID-19 pandemic, and in July 2022, as access to the village was prohibited during the pandemic. The pigs were located outside the residences, being kept in closed pigsties with restricted access to areas close to the houses. Of the 18 samples collected, 44.4% were positive for *Ascaris* spp. Table 2.

**Table 2.** Total and species-specific prevalence of intestinal protozoa and helminths in pigs from the village of Santa Rosa do Ocoy, municipality of São Miguel do Iguaçu, state of Paraná, southern Brazil (n=18). Samples collected in March 2020 and July 2022.

| Species | No. positive samples | Prevalence (%) |
| --- | --- | --- |
| **Total** | 10 | 55.6 |
| *Ascaris* spp. | 8 | 44.4 |
| *Balantidium coli* | 2 | 11.1 |
| *Entamoeba* sp. | 1 | 5.6 |
| *Trichuris trichiura* | 1 | 5.6 |
| Hookworms (larva) | 1 | 5.6 |

Of the 68 soil samples collected, 52 (76.5%) were positive for at least one parasite species, the majority with zoonotic potential for infection in humans. Six samples (8.9%) were positive for *Ascaris* spp. Table 3.

**Table 3.**
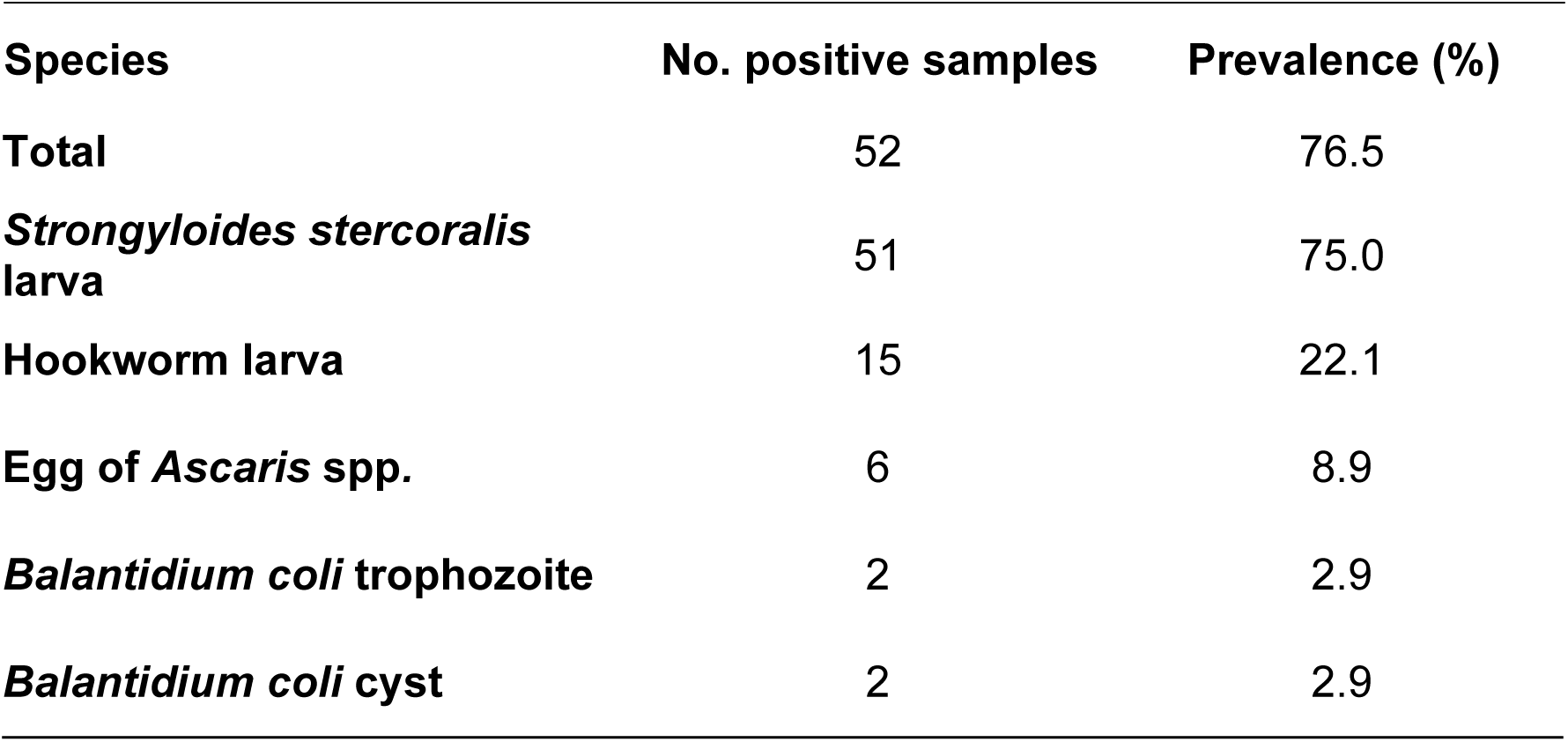
Total and species-specific prevalence of parasitic forms found in peridomiciliary soil samples from the village of Santa Rosa do Ocoy, municipality of São Miguel do Iguaçu, state of Paraná, southern Brazil (n=68). Samples collected throughout 2019 and 2021.

Sediments of fecal and soil samples, obtained using the Ritchie method adapted by Bezagio et al. [29], were sonicated to rupture the *Ascaris* spp. egg membrane (Fig 3), thus favoring DNA extraction, as described below.

**Fig 3.**
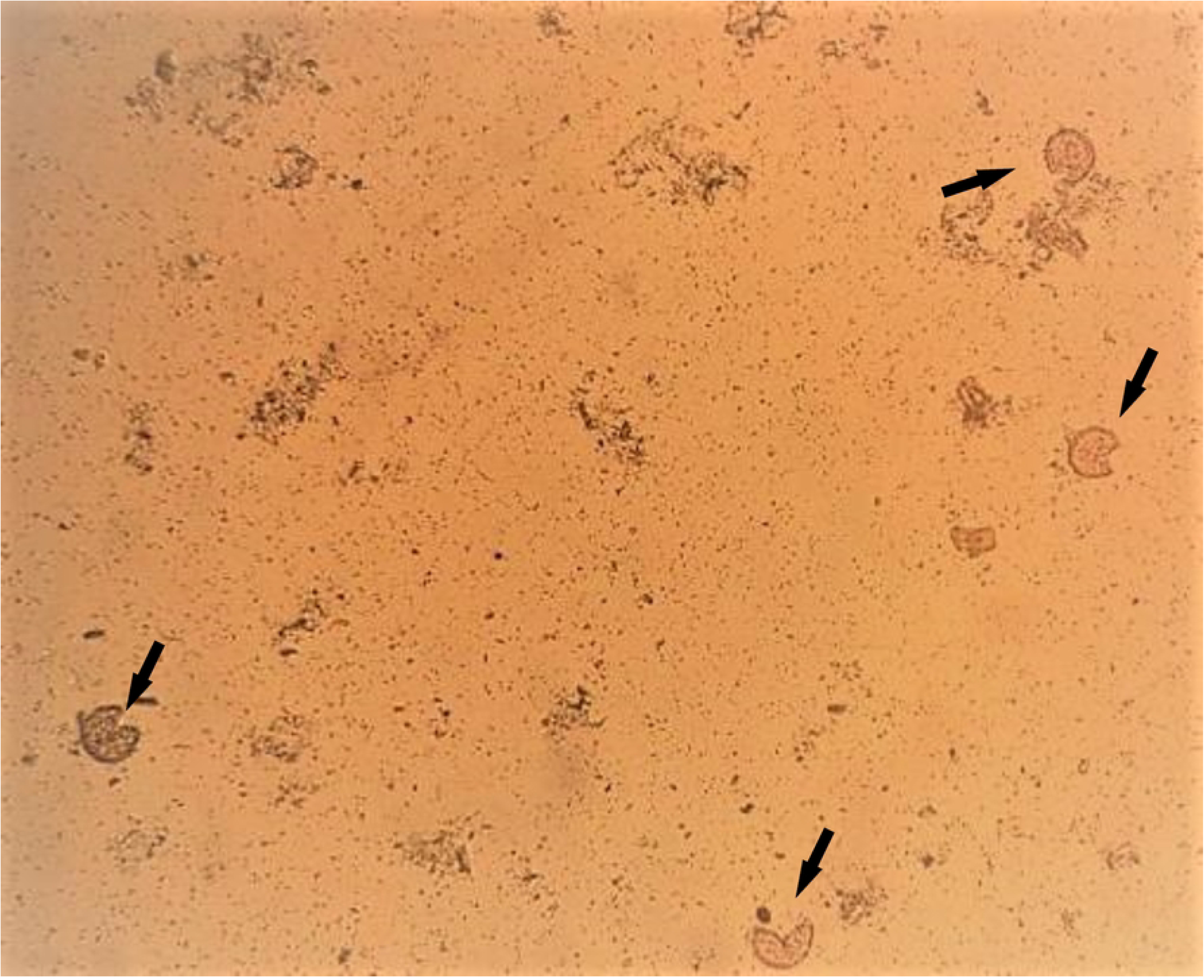
Microphotograph of fecal sample sediment showing eggs of *Ascaris* spp. ruptured. (arrows) after sonication at 50 Hz for 30 s at 4°C, repeated 4 times, with a 1 min interval between cycles. 100× magnification. Source: the author

Three DNA extraction/purification methods were evaluated using a known *A. lumbricoides*-positive sample and all methods appeared to be efficient. Subtle differences were observed in the intensities of the *ITS-1* locus band of the extracted samples following PCR (Fig 4). Extraction with the PureLink^®^ PCR Purification Kit (Invitrogen, Carlsbad, CA, USA) preceded by sample sonication resulted in a slightly more intense band than the other two protocols tested, and thus, this method was chosen for the DNA extraction of samples collected in this study (Fig 4).

**Fig 4.**
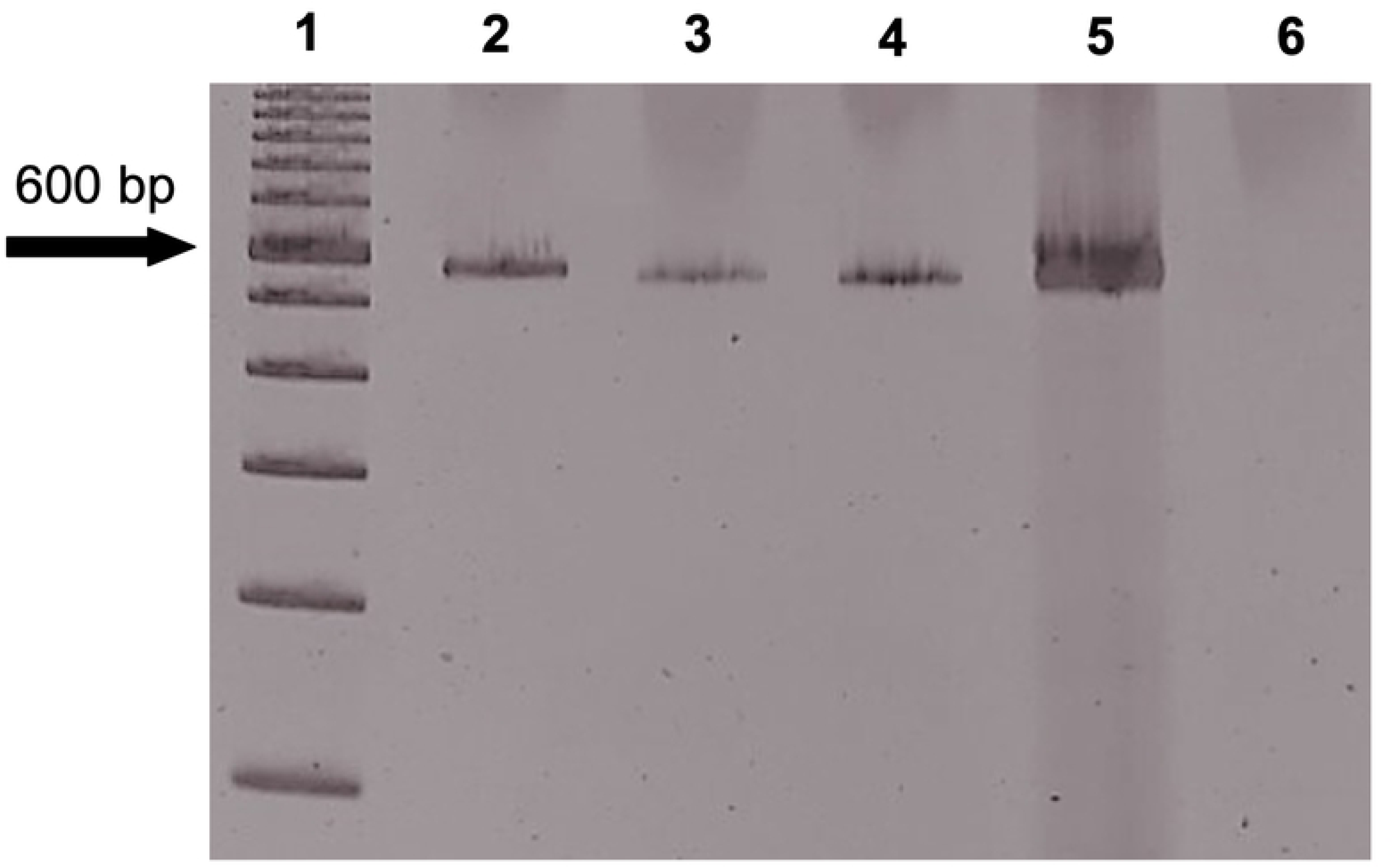
A 5% polyacrylamide gel showing bands of approximately 580 bp characteristic of the *ITS-1* locus of *Ascaris* spp. 1. DNA ladder; 2. *A. lumbricoides*-positive laboratory sample extracted with the PureLink^®^ PCR Purification Kit after sonication (50 Hz for 30 sec at 4°C repeated 4 times with 1 min interval between cycles); 3. *A. lumbricoides*-positive laboratory sample extracted with the PureLink^®^ PCR Purification Kit without sonication; 4. *A. lumbricoides*-positive laboratory sample extracted with the QIAmp^®^ DNA Stool Mini Kit after sonication; 5. positive control (*A. lumbricoides* DNA); and 6. negative control (PCR reaction without DNA).

The sediments of the collected samples (human and pig feces and soil) were submitted to DNA extraction followed by PCR amplification of the *ITS-1* locus. Genotyping was then carried out using the PCR products from the *ITS-1* amplification reaction to verify the presence of *A. lumbricoides* and *A. suum*. Table 4 shows the results of microscopy analysis and the *ITS-1* genotyping in the three sample types. The DNA of *Ascaris* spp. was amplified from all samples that were positive in the microscopy analysis. Twice as many samples that were negative in the microscopy were randomly chosen to undergo the amplification reaction for validation, and all were negative.

**Table 4.** Parasitological and molecular detection of *Ascaris* spp. in fecal samples from indigenous Guarani schoolchildren and pigs, as well as soil samples, using optical microscopy and PCR genotyping of the *ITS-1* locus. Samples were collected from the village of Santa Rosa do Ocoy, municipality of São Miguel do Iguaçu, state of Paraná, southern Brazil.

| Sample | Origin | Microscopy | ITS-1<br>PCR | Species |
| --- | --- | --- | --- | --- |
| H05 | Human | - | - |  |
| H06 | Human | - | - |  |
| H07 | Human | + | + | AI |
| H09 | Human | - | - |  |
| H23 | Human | + | + | AI |
| H27 | Human | - | - |  |
| H31 | Human | - | - |  |
| H32 | Human | + | + | AI |
| H36 | Human | - | - |  |
| H41 | Human | - | - |  |
| H50 | Human | - | - |  |
| H61 | Human | - | - |  |
| H66 | Human | - | - |  |
| H70 | Human | - | - |  |
| H71 | Human | - | - |  |
| H78 | Human | - | - |  |
| H79 | Human | + | + | AI/As |
| H80 | Human | + | + | AI |
| H81 | Human | - | - |  |
| H82 | Human | + | + | AI/As |
| H83 | Human | + | + | AI/As |
| P01 | Pig | - | - |  |
| P02 | Pig | - | - |  |
| P03 | Pig | - | - |  |
| P04 | Pig | - | - |  |
| P05 | Pig | + | + | As |
| P06 | Pig | + | + | As |
| P07 | Pig | + | + | As |
| P08 | Pig | - | - |  |
| P09 | Pig | + | + | As |
| P10 | Pig | - | - |  |
| P11 | Pig | - | - |  |
| P12 | Pig | - | - |  |
| P13 | Pig | + | + | As |
| P14 | Pig | + | + | As |
| P15 | Pig | - | - |  |
| P16 | Pig | + | + | As |
| P17 | Pig | - | - |  |
| P18 | Pig | + | + | AI/As |
| S1I | Ground | + | + | As |
| S2I | Ground | - | - |  |
| S3I | Ground | - | - |  |
| S5I | Ground | - | - |  |
| S10I | Ground | - | - |  |
| S12I | Ground | - | - |  |
| S15I | Ground | + | + | AI/As |
| S16I | Ground | - | - |  |
| S01P | Ground | - | - |  |
| S02P | Ground | + | + | AI |
| S03P | Ground | - | - |  |
| S05P | Ground | - | - |  |
| S07P | Ground | + | + | AI/As |
| S10P | Ground | - | - |  |
| S11P | Ground | + | + | As |
| S12P | Ground | + | + | AI |
| S14P | Ground | - | - |  |

+ Positive; - Negative; AI, *Ascaris lumbricoides*; As, *Ascaris suum*.

Of the human fecal samples that were positive in the parasitological examination and exhibited amplification of the *ITS-1 locus* (7/83), 57.1% (4/7) presented a band of ∼396 bp, characteristic of *A. lumbricoides*, and 42.9% (3/7) showed a ∼396 bp band together with a band of ∼176 bp, characteristic of *A. suum*. The presence of both bands indicated a mixed infection of the two *Ascaris* species in humans. Among the positive pig fecal samples (8/18), 87.5% (7/8) presented the ∼178 bp band and 1/8 (12.5%) showed both bands, suggesting mixed infection in these animals. Of the *Ascaris-*positive soil samples (6/68), 33.3% (2/6) showed the *A. lumbricoides* characteristic band, 33.3% (2/6) showed the *A. suum* characteristic band, and 33.3% (2/6) showed both bands. Some representative samples of these results are presented in Fig 5.

**Fig 5.**
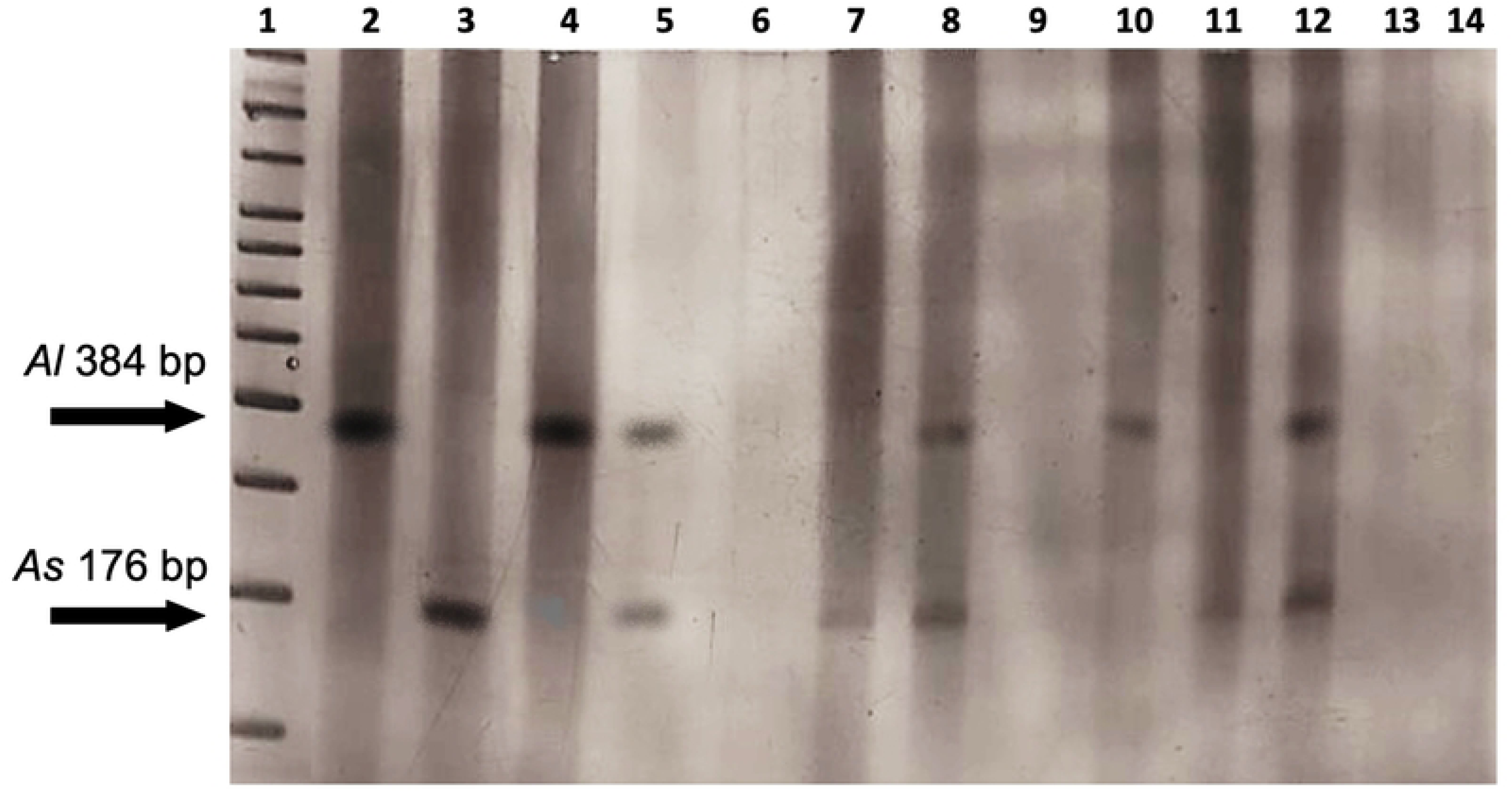
A 5% polyacrylamide gel showing the bands resulting from the *ITS-1* locus genotyping PCRs of human and pig fecal samples and soil samples. The band for *A. lumbricoides* is ∼384 bp and for *A. suum* is ∼176 bp. 1: DNA ladder; 2: positive control for *A. lumbricoides*; 3: positive control for *A. suum*; 4 and 5: human feces samples that were positive in the parasitological examination; 6: human feces samples that were negative in the parasitological examination; 7 and 8: pig feces samples that were positive in the parasitological examination; 9: pig feces samples that were negative in the parasitological examination; 10, 11, and 12: soil samples that were positive in the parasitological examination; 13: soil sample that was negative in the parasitological examination; 14: negative control.

## Discussion

This is the first molecular epidemiology study to determine the prevalence of *Ascaris* spp. in an indigenous community of Brazil, genetically and culturally distinct from the surrounding society, and to evaluate the circulation of this pathogen between humans, pigs, and the environment (soil).

Human ascariasis is a serious public health problem in developing countries [18]. The biological cycle of the parasite is favored by poor basic sanitation, poor hygiene practices, and a population living in poverty, which, together with climatic conditions, contribute to high levels of prevalence, especially in school-age children [1,3,4,32,33].

In this study, the highest prevalence of *Ascaris* spp. was recorded in pigs (44.4%), followed by the soil (8.8%) and, finally, in humans (8.4%). The low prevalence of *Ascaris* spp. observed in schoolchildren from this Guarani village does not correspond to the reality of many other Indigenous Lands in Brazil; in a study conducted in the state of Minas Gerais, southeastern Brazil, the infection rate was reportedly 34.4% [34], while a 63.5% infection rate was reported for another region of Paraná [24]. This divergence could be the result of periodic antiparasitic treatment and, to a lesser extent, to housing improvements implemented by the government in this region in recent years [19,24,34,35].

Although the prevalence of soil-transmitted helminths, such as *Ascaris* spp., *Trichuris trichiura*, and hookworm, and intestinal helminths in the schoolchildren of the village was generally low (32.5% compared to 74.7% for intestinal protozoa), *Hymenolepis nana* was the most prevalent helminth in this population. Similar prevalence rates of *H. nana* have been reported for other Indigenous Lands of Paraná, inhabited by Kaingáng indigenous people, where similar prophylactic measures were adopted [24].

The epidemiological picture of human ascariasis around the world has shown some considerable changes, with increasing reports of this parasitic disease in developed countries, where cases of infection in humans were low or non-existent [36,37]. While *A. lumbricoides* typically affects humans and *A. suum* infects pigs [10], when human infection during travel or residence in endemic areas is ruled out, contamination with fecal material from pigs has been identified as the main source of infection [38]. Studies have shown that non-endemic regions usually present high levels of cross-infection, with humans being infected by *Ascaris* spp. derived from pigs, as well as a high percentage of hybrids, and thus, ascariasis has been characterized as a zoonosis in these regions [36, 39–41]. These reports strongly point to the zoonotic potential of *A. suum* [6,38,42], and the ability of *A. suum* to develop infection in experimentally infected humans has already been demonstrated [43].

The methods routinely used for the diagnosis of ascariasis do not allow identification at the species level, owing to the indistinguishable morphology of the eggs between species. The diagnosis is therefore inferred based on the host species under analysis and the epidemiology of the region [44–46]. However, given the changing epidemiological picture, molecular techniques that allow the identification of *Ascaris* species from the DNA of a single egg have been developed. These typically require the use of three primers in the same reaction, in which two primers are species-specific (one for each allele) and one primer is common [31,47,48].

To perform these molecular analyses, DNA must first be extracted from the sample. Three DNA extraction/purification procedures were evaluated in the current study. As *Ascaris* spp. eggs have three membranes that protect from environmental changes [49], sonication of the samples prior to DNA extraction appeared to favor the extraction process [50] and was performed prior to extraction with the PureLink PCR Purification^®^ Kit.

It was possible to identify and differentiate the species of *Ascaris* through the molecular analysis of the *ITS-1* locus of samples of human and pig feces, as well as soil, and thus, the occurrence of cross-infection in the village could be investigated. In this study, 42.9% of positive human fecal samples presented both *A. lumbricoides* and *A. suum*, suggesting the occurrence of swine-human transmission of *Ascaris* in a high proportion of infected individuals. This is different from other studies which have demonstrated that, in endemic regions, cases of human infection are the result of human-to-human transmission [14, 39]. The description of 54 dominant haplotypes and genotypes in *Ascaris* originating from a given host reveal a host affiliation in sympatric populations of endemic regions [50–53]. Therefore, it is crucial to understand the distribution of haplotypes and frequency of hybrids in the *Ascaris* population in a given region [41,53–56].

The *ITS-1* locus was used as the molecular target of choice in this study because it is a region with multiple copies in the parasite genome and has well-characterized single nucleotide polymorphisms between the two species. At nucleotide position 133, a guanine is present in the *A. lumbricoides* sequence and a cytosine in the *A. suum* sequence; at position 246 a thymine is present in the *A. lumbricoides* sequence and an adenine in the *A. suum* sequence; and at position 323 an adenine is present in *A. lumbricoides* and a guanine in *A. suum* [14,36,57].

The results of the molecular analyses confirmed the results of the parasitological microscopy analyses and showed that the samples analyzed in this study demonstrated a pattern of transmission between host species. This corroborates the results of analyses carried out by other research groups who, using mitochondrial (mt)DNA and microsatellite markers, analyzed the sharing of *Ascaris* haplotypes and genotypes between Brazilian human and swine populations, indicating that cross-infection is occurring in some regions [11,53,56].

In our study, PCR for the *ITS-1* region did not detect *Ascaris* DNA in any of the samples that were negative in the parasitological examination. In other words, the two techniques used in the examination of the biological materials, light microscopy and PCR, showed high sensitivity, that is, the ability to detect the parasite. However, the molecular technique has an advantage over parasitological techniques by enabling the identification of the species/genotype of the parasite present in the sample.

In other Indigenous Lands of Paraná inhabited by other ethnic groups, such as the Kaingáng, where pigs are raised freely, in an extensive system, the soil constitutes an important source of parasitic contamination since, as already demonstrated, the same species of intestinal parasites can be found in human feces and soil [24,40,58]. Differently, in the Guarani indigenous village of the present study, pigs are raised in a non-extensive, confined regime. However, as was observed *in situ* by researchers, pigs occasionally escape from the pens, being able to defecate in areas where people circulate, and contaminate the soil with species of parasites with zoonotic potential.

Despite the low rate of contamination by *Ascaris* spp. found in this study when compared to the rates reported for studies of other Indigenous Lands [24,33,40,59], the human-soil-swine transmission link was demonstrated by genotyping and the mixed infection findings in humans and pigs, and the contamination of soil by both species.

### Conclusions

This study reveals cross-infection by the two nematode species, *A. lumbricoides* and *A. suum*, in human and swine hosts from a Guarani indigenous village in southern Brazil. In addition to the hosts, the soil was contaminated with both species of *Ascaris*. Pure and genetically mixed or hybrid samples were observed in both hosts and in the soil, confirming that is a source of infection for human and animal populations.

Given the cross-infection in the village, which confirms the zoonotic nature of ascariasis, more effective and targeted control measures must be implemented, such as better containment of pigs to prevent access to human feces, in order to reduce the infection rates of *Ascaris* spp.

The molecular analysis of *Ascaris* in humans and pigs can contribute to the development of control measures, and future studies should be carried out in this and other sympatric areas to further enhance our knowledge about the transmission dynamics of this parasite.

## Data Availability

Our article does not report data and the data availability policy is not applicable to the article.

## Acknowledgments

The authors would like to express their gratitude to Lilian Maria Rodrigues and Bonifácia Rero Takua Alves for their valuable assistance in recruiting participants, Eloiza Cristina Martelli and Guilherme Martins Boeira in collecting the soil and pigs’ feces for this study.

